# Modelling the relative benefits of using the measles vaccine outside cold chain for outbreak response

**DOI:** 10.1101/2021.05.19.21257492

**Authors:** James M. Azam, Barbara Saitta, Kimberly Bonner, Matthew J. Ferrari, Juliet R.C. Pulliam

**Author notes:** Corresponding author, Corresponding author physical address: SACEMA, 19 Jonkershoek Road, Stellenbosch 7600. denotes joint senior authorship (Matthew J. Ferrari, Juliet R.C. Pulliam).

## Abstract

**Introduction:** Rapid outbreak response vaccination is a strategy for measles control and elimination. Measles vaccines must be stored and transported within a specified temperature range, but this can present significant challenges when targeting remote populations. Measles vaccine licensure for delivery outside cold chain (OCC) could provide more vaccine transport/storage space without ice packs, and a solution to shorten response times. However, due to vaccine safety and wastage considerations, the OCC strategy will require other operational changes, potentially including the use of 1-dose (monodose) instead of 10-dose vials, requiring larger transport/storage equipment currently achieved with 10-dose vials. These trade-offs require quantitative comparisons of vaccine delivery options to evaluate their relative benefits.

**Methods:** We developed a modelling framework combining elements of the vaccine supply chain - cold chain, vial, team, and transport equipment types - with a measles transmission dynamics model to compare vaccine delivery options. We compared 10 strategies resulting from combinations of the vaccine supply elements and grouped into three main classes: OCC, partial cold chain (PCC), and full cold chain (FCC). For each strategy, we explored a campaign with 20 teams sequentially targeting 5 locations with 100,000 individuals each. We characterised the time needed to freeze ice packs and complete the campaign (campaign duration), vaccination coverage, and cases averted, assuming a fixed pre-deployment delay before campaign commencement. We performed sensitivity analyses of the pre-deployment delay, population sizes, and two team allocation schemes.

**Results:** The OCC, PCC, and FCC strategies achieve campaign durations of 50, 51, and 52 days, respectively. Nine of the ten strategies can achieve a vaccination coverage of 80%, and OCC averts the most cases.

**Discussion:** The OCC strategy, therefore, presents improved operational and epidemiological outcomes relative to current practice and the other options considered.

## Introduction

Measles outbreaks have surged worldwide and more so in low-income countries, causing severe morbidity and high levels of mortality [1]. Measles is prevented by a safe, cheap and effective vaccine [2]. A high level of vaccination coverage (> 95%) is required to successfully prevent outbreaks. However, there is a stagnation in local-level vaccination coverage in many areas, leading to recurrent outbreaks [1]. In such settings, outbreak response vaccination is a proven strategy for reducing the impact of the outbreaks [3]. The potential impact of an outbreak response vaccination campaign depends on the coverage achieved and the speed with which the campaign is completed relative to outbreak spread. There is, therefore, a desperate need to improve vaccine delivery strategies during outbreak response campaigns [4].

The measles vaccine, being a biological product, has been licensed for storage and use in the cold chain within a recommended temperature range of 2-8°C [2]. This temperature requirement, along with the need to use freezers, refrigerators, and ice packs, and special transport and storage equipment to maintain it, is extremely challenging in settings with poor road and health infrastructure [5]. These challenges limit the ability to achieve high vaccination coverage rapidly [4,5].

A recent study found that the measles vaccine remains stable and efficacious for up to 6 days when exposed to ambient temperatures up to 37°C and 2 days when exposed to 40°C, meeting the World Health Organisation’s definition of Extended Controlled Temperature Conditions (ECTC) [6]. ECTC refers to *“approved short-term temperature conditions, above those defined for long-term storage, transportation and use, for a given product immediately prior to administration*” (WHO, 2015). Consequently, the measles vaccine can, in principle, be used outside cold chain (OCC), if prequalified and relabelled under ECTC. MenAfriVac^®^, a Meningitis A vaccine, and Gardasil, an HPV vaccine have been labelled for use under ECTC, with growing evidence of significant economic and public health benefits [9–14]. Here, we evaluate the potential logistical and epidemiological benefits of using the measles vaccine OCC for outbreak response campaigns.

Using the measles vaccine OCC under ECTC could have many advantages. The vaccines will be stored and transported in the cold chain to field bases where the final preparations will be done for campaigns. However, teams will not need ice packs to commence the campaign at remote distribution points. This will therefore eliminate the time spent in freezing ice packs before commencing the campaign. This gain in lead time will enable quick commencement, which coupled with a high coverage, can have a significant impact on the outbreak size [15]. The reduction in ice pack needs may also lead to a more efficient use of the available transport capacity.

Our objective was to compare the operational and epidemiological benefits of potential vaccine delivery options, including using the measles vaccine in OCC under ECTC, for outbreak response vaccination. We achieved this by developing a modelling framework that allowed us to evaluate the proposed alternative strategies in terms of three metrics: (a) campaign duration, (b) achieved vaccination coverage, and (c) cases averted.

## Material and methods

We developed a modelling framework that combines a vaccine supply chain model with a Susceptible-Exposed-Infected-Recovered (SEIR) model to characterize the preparation time required to commence and complete a vaccination campaign (campaign duration), the vaccination coverage achieved, and the dynamics of the resulting outbreak happening in parallel (). The individual components of the framework are discussed below.

In formulating a suitable modelling framework to achieve our objective, we used an approach that aligns with the elements of a structured decision framework for evaluating interventions: a clear question/objective, a clear definition of the interventions, the models and modelling approaches for assessing the interventions, and the sources of uncertainties in the decision or recommendation [16,17].

The SEIR modelling framework typically does not explicitly capture the vaccine supply chain processes involved in an outbreak response campaign. Hence, faced with evaluating strategies involving the use of two vial types (10-dose vs. monodose), with or without the full cold chain, and considering different transport and storage equipment with capacity constraints, we developed a modelling framework that could capture these additional elements. We therefore extended the traditional SEIR framework to include a vaccine supply chain model that captured the requirements of the two vial types, cold chain, and equipment and team types along with their relevant characteristics (Fig. 1). The individual models making up the framework will be discussed in later sections.

**Fig. 1.**
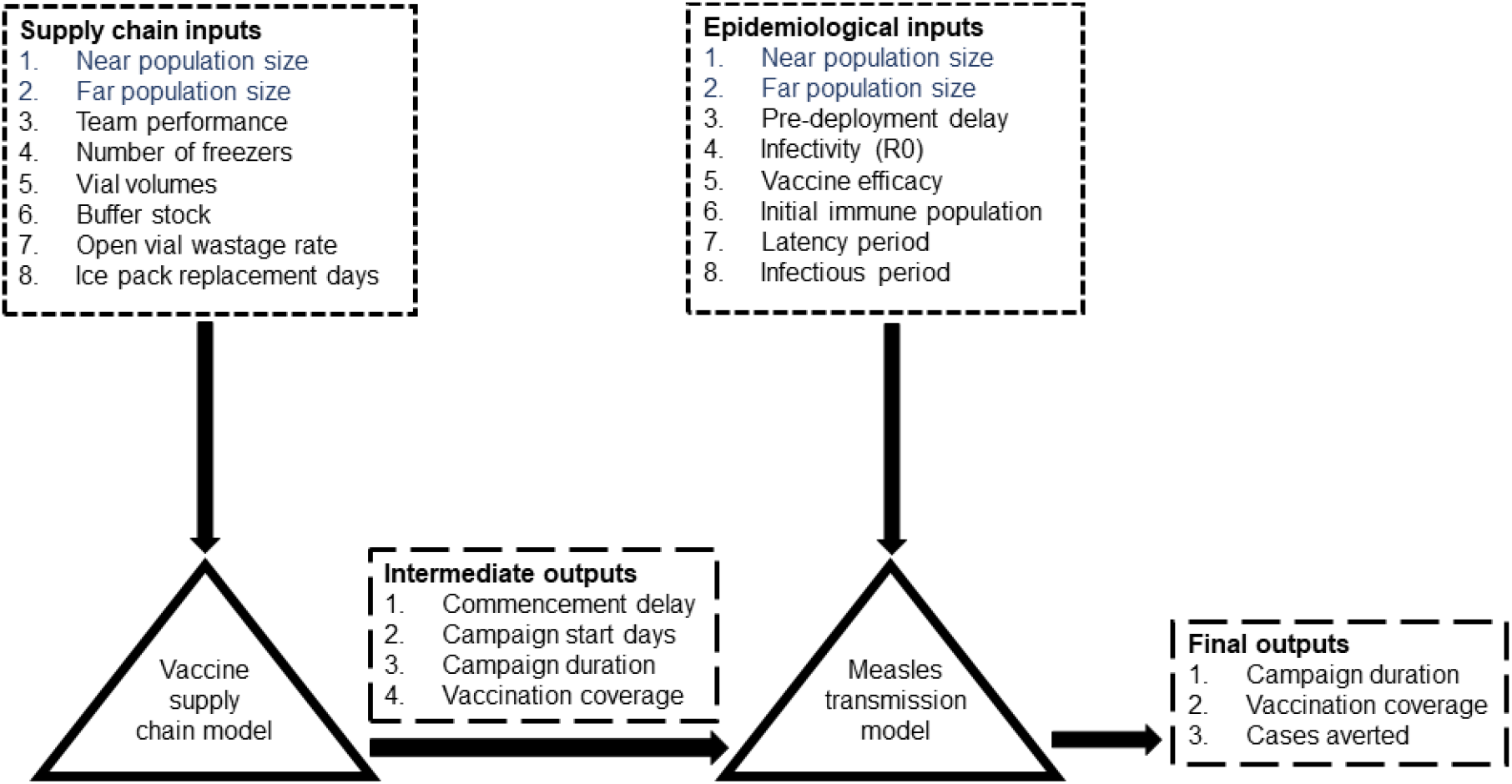
Schematic of the modelling framework. The vaccine supply chain model and measles transmission model form the core of the framework. The two models both have unique inputs (black) and shared inputs (blue). Inputs and outputs are indicated with the black arrows. The supply chain model is used to estimate the delay to commence a campaign, campaign start days at each location, the campaign duration, and vaccination coverage, which are used as inputs to the epidemiological model. The transmission model simulates the transmission dynamics and is used to calculate the cases averted from each strategy. The strategies can then be compared based on campaign duration, vaccination coverage, and cases averted.

### Elements of a typical outbreak response vaccination campaign

During outbreak response vaccination campaigns, measles vaccines are stored and transported in passive cold chain equipment [18]. All passive cold chain equipment depends on ice packs to keep the vaccines cold. Additionally, campaigns require active cold chain equipment, for example freezers and refrigerators, which are less mobile and require a continuous power supply to freeze ice packs for transporting the vaccines [18].

RCW25 cold boxes and vaccine carriers are two of the most commonly used types of passive cold chain equipment [18]. RCW25 cold boxes can transport up to 6 times more doses of vaccine than the vaccine carrier but also require more ice packs due to their large internal volume. The RCW25s are often used to store vaccines intermediately during a vaccination session while small quantities of the vaccines are transferred into the vaccine carrier for preparation and administration.

The measles vaccine comes in several vial presentations: monodose (1-dose) and multidose (2-, 5-, or 10-doses per volume). Multidose vials require less passive cold chain compared to monodose vials for the same number of doses. Monodose vials, on the other hand, reduce open vial wastage, where opened multidose vials are discarded if they are not completely used after a certain period [19]. Open vial wastage has the potential to affect vaccine stock during a campaign and negatively impact on the vaccination coverage (proportion of the total population vaccinated).

### The modelled campaign

A typical vaccine supply chain for an outbreak response campaign comprises the levels from top to bottom as shown in Fig. 2. The number of levels and facilities per level may differ based on the context. The OCC strategy can only be applied in the grey-shaded area due to the time constraints activated when the vaccine is exposed to ambient temperatures; we thus focus our model on this part of the vaccine supply chain as measles vaccine is only stable for 2-6 days OCC.

**Fig. 2.**
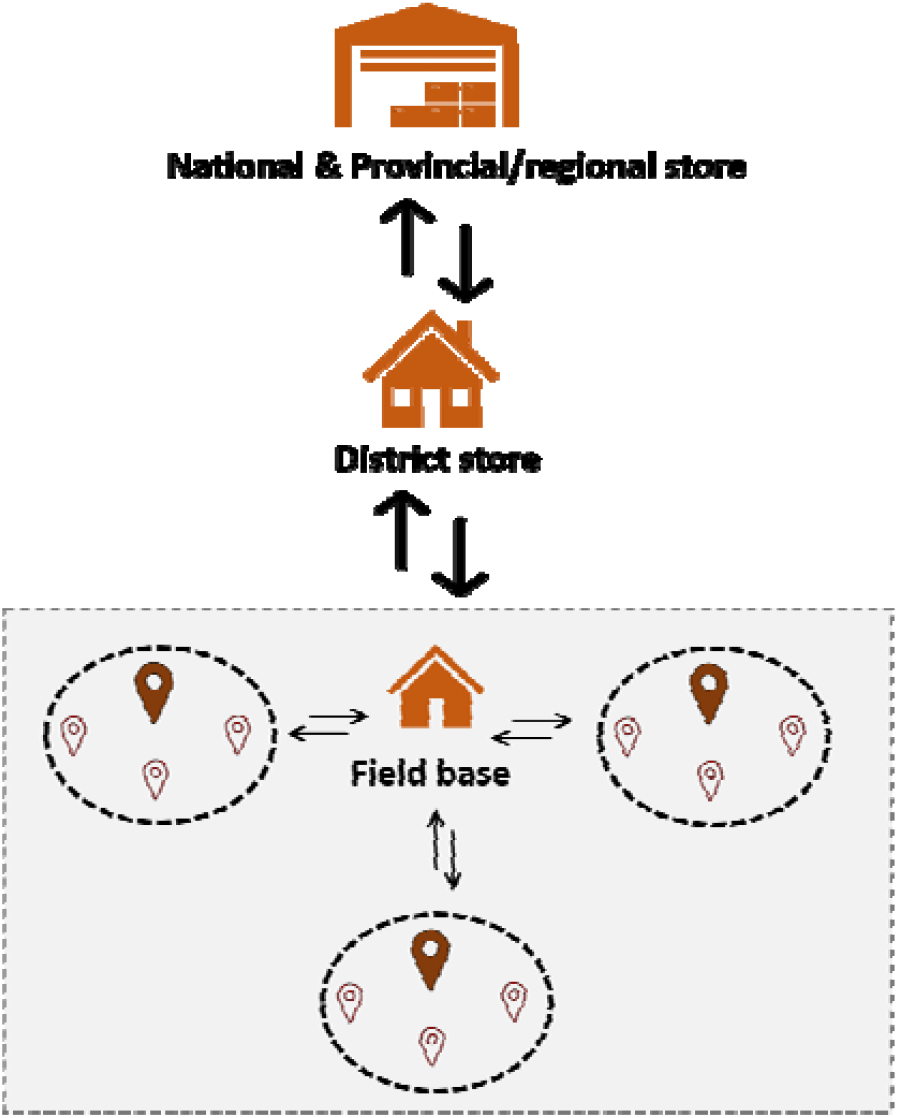
A generic vaccine supply chain. The number of supply chain levels could differ based on the context. Moreover, there are usually several interactions (arrows) between the various supply chain levels. The shaded region represents the level where the OCC strategy could be implemented to harness its full benefits. Only this region was modelled in this study. In the shaded level, a field base is set up as a hub to serve the target locations (dashed ovals). From the field base, the teams (fixed post and mobile teams) are prepared and dispatched to the target locations in serial. At each location, the fixed post teams set up a vaccination site (brown filled icons within dashed ovals) to target the near population, and the mobile teams travel to the far populations (hollow icons with brown outline within dashed ovals).

We described an outbreak response campaign (grey rectangle in Fig. 2) as an immunization activity targeting multiple locations in serial order.

Within each location, we categorised the populations as either “near” or “far”. We considered near populations to be those in the urban areas who are easily accessible with a stationary (“fixed post”) team. Far populations on the other hand are those situated in remote, hard-to-reach locations that can only be reached by a mobile team that transports the vaccines either with a vaccine carrier or an RCW25. For traditional cold chain campaigns, an RCW25 is rarely used as transport equipment without an accompanying vaccine carrier.

For OCC, we consider campaigns that do not require passive cold chain, so the use of RCW25s and vaccine carriers (which are designed to maintain temperature using ice packs) is not required. However, since these are standard equipment and the volumes are known, we consider these as the equipment that would be used to transport vaccine doses OCC. We assume that the volume that would normally be allocated to ice packs is available to carry additional doses in the OCC strategies.

We defined two team types to serve the two population categories in a location: fixed post teams serve the near populations and mobile teams serve the far populations. During a campaign in each location, the fixed post teams set up vaccination sites to administer the vaccines. Fixed post teams expect the near population to show up for vaccination. In contrast, the mobile teams actively commute to the far populations to administer the vaccines.

For a complete campaign (Fig. 2**Error! Reference source not found**.), we set up one field base to serve as a hub for preparing and dispatching the two team types to the vaccination sites in 5 locations. We set up the supply chain model to account for the time it takes to undertake two activities at the field base: (a) freezing of ice packs for transporting the vaccines (b) dispatching the fixed post and mobile teams together as one unit. All these activities contribute to the time between the initiation of a campaign and the beginning of vaccination.

We assumed that the campaign is conducted by the two team types in parallel in each location but sequentially between the locations. Furthermore, we assumed that the fixed post and mobile teams work independently at each location but move together to the next location so that the time spent in each location is determined by the slower of the two team types.

In the model, the teams move from one location to the next when the campaign has reached its stopping condition in the previous location. The stopping condition for each location is determined as the minimum of two factors: (a) a fixed campaign duration allowable per location and (b) the time required to achieve 100% vaccination coverage in the location. The stopping condition here is informed by real campaign situations where governments may restrict the vaccination campaigns to a specified duration [4].

### Supply chain model

The supply chain model was formulated to estimate three strategy-dependent outcomes: namely, the commencement delay, campaign duration, and vaccination coverage.

Outbreak response vaccination teams can only be dispatched to commence the campaign when they have enough ice packs to transport the vaccines. We therefore formulated the supply chain model to calculate the time required to freeze ice packs to dispatch teams, which we term the commencement delay. The time to freeze ice packs serves as an input to the epidemiological model.

Assuming there are *L* locations, each with population sizes categorised into near, *N*_*near*_, and far, *N*_*far*_, we can write the total target population size, *N*, as

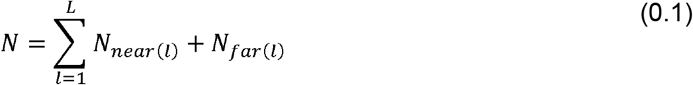

where *I* represents a single location and *1 ≤I ≤ L*. Therefore, each location *I* has a total target population of size *N*_*i*_, which we assumed is known.

Using the total population size of each location, *N*_*l*_, we calculated how many doses, *Dl*, of vaccine are needed for the campaign in the first location, *D*_*l*_, the second, *D*_*2*_, and so on. We adjusted the number of doses with a buffer stock, *b*, defined as a proportion relative to *N*_*l*_, to account for damaged vials and other unexpected demand. Also, we assumed an open vial wastage rate, *w*, associated with each vial type. This kind of adjustment is common practice in vaccine inventory calculations [20].

Therefore, the number doses required for the campaign in location, *I*, is given as

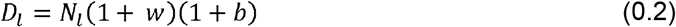

The amount of passive cold chain equipment needed for the campaign depends on the number of each team type to be dispatched. We assumed based on the Médecins Sans Frontières (MSF) measles outbreak management guide[21] that each fixed post has two fixed post teams and required 1 RCW25 and 2 vaccine carriers.

Based on the Médecins Sans Frontières (MSF) measles outbreak management guide, the number of ice packs needed for use with the transport equipment depends on the type of transport equipment, ambient temperature, and the number of days after which to replace the ice packs [21]. Ice pack replacement is necessary because the ambient temperature eventually thaws them. We assumed that the ambient temperature would be below 40°C and that the ice packs required for both equipment types would be replaced each day. Therefore, each vaccine carrier and RCW25 required 6 small ice packs (0.4L) and 12 large ice packs (0.6L) respectively.

We denote the total number of small and large ice packs needed, based on the number of teams, as Ø_*small*_ and Ø_*large*_ respectively. We can estimate the time needed to freeze the ice packs given we know the number of freezers and their ice pack freezing rates. For this study, we assumed that the field base uses the MF314 type of freezer, which is the biggest of the freezers often used for MSF’s outbreak response activities [21]. We have provided its specifications in Table S1 in the supplementary material.

Given the number of MF314 freezers, *f*, at the field base and the number of ice packs that can be frozen per day in one freezer for the small and large ice packs, *r*_*small*_ and *r*_*large*_ respectively, we compute the time it takes to freeze the ice packs, *t*_*ice packs*_, as

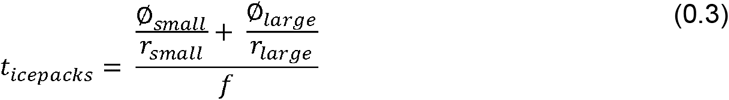

We defined the team-days, *d*, as the number of days that would be required in any location by a single team to achieve 100% coverage. We used the team-days to determine how long a campaign would take in each location, assuming a constraint on the campaign duration allowed at each location, and a specified number of available teams. The campaign duration at each location was, therefore, the minimum of the team-days per team type divided by the number of teams, and the maximum campaign duration. If the number of days required to reach 100% coverage was higher than the campaign duration allowed, the vaccination coverage achieved in that location was <100%.

The team-days is a function of the effective doses and the team performance, *P*. The effective doses a team can administer on a given day of the campaign is *v\** (1*-w*,), where *v* is the transport capacity, and *w* is the open vial wastage rate. The transport capacity, *v*, is determined by the passive cold chain equipment (vaccine carrier or RCW25 or both) used, and whether the strategy requires ice packs or not (see Table S2). The team-days formula for fixed post, *d*_*fixed*_, and mobile teams, *d*_*mobile*_, are therefore given as

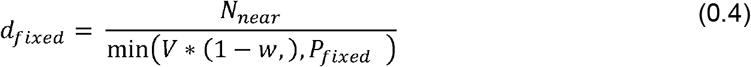

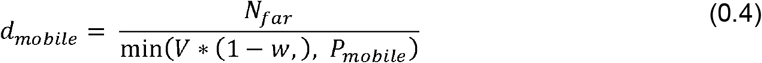

If there is more than one team working in parallel in a location, the team-days can be allocated between teams to reduce the campaign duration. Let *T*_*fixed*_ (*I*) and *T*_*mobile*_ (*I*) be the number of fixed post and mobile teams dispatched on a campaign to location, *I*, with near and far target populations of sizes, *N*_*near*_(*I*) and *N*_*far*_(*I*). Given that a single fixed post team needs *d*_*fixed*_ (*I*) team-days and a mobile team needs *d*_*mobile*_(*I*) team-days in this location to achieve 100% coverage, then the per-fixed-team campaign duration assuming all the *T*_*fixed*_ (*I*) fixed teams have the same work rate is given as

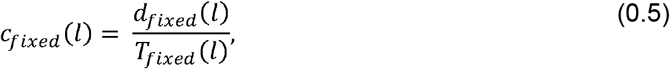

while that for mobile team is given by

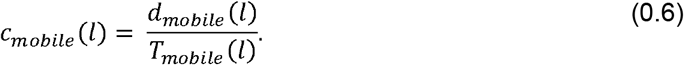

Given that the campaign at any location, *I*, is constrained by a campaign duration *C*_*max*_ (*I*), and the campaign duration in any location is determined by the slower of the two team types, then the campaign duration in location *I* is given as

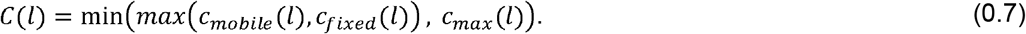

Recall that the campaign commencement delay was derived in (0.3). Combining (0.3) and (0.7), we obtain the campaign duration of a sequential campaign in *L* locations as

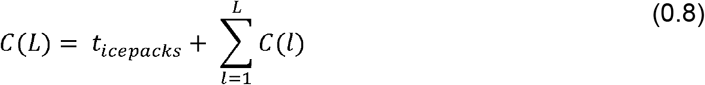

We assumed that at each location,*I*, the teams were either constrained by the effective doses, which accounts for open vial wastage, or team performance. Therefore, the vaccination coverage of the two population types per location, *v*_*p*_, where *p∈* {*near, far*}, and team type *J ∈*{*fixed, mubiIe}*, was calculated as

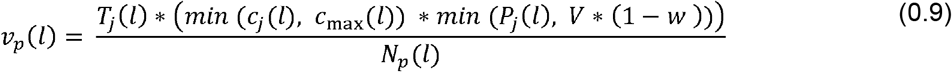

Hence, the vaccination coverage across all *L* locations is given as

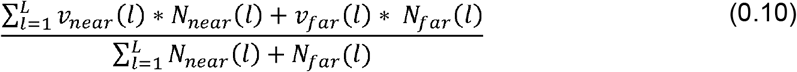

### Epidemiological model

We modelled the transmission dynamics at each location with a deterministic continuous-time compartmental model [22]. This model used the main supply chain outputs - commencement delay, campaign duration, and vaccination coverage by location - as its inputs (Fig. 1). We also assumed a fixed pre-deployment delay, *φ*,to account for the time until vaccines arrive at the field base.

We categorised the near and far populations in each location into four disease states: Susceptible (S), Exposed but not yet infectious (E), Infectious (I), and Recovered (R). The system of ordinary differential equations describing the model is as follows

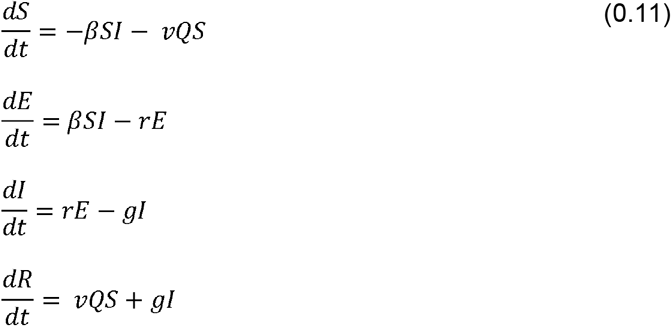

Where *β, 1/ r,1/g, and v* represent respectively the transmission rate, latency period, infectious period, and vaccine efficacy. Additionally, *Q* is the vaccination rate obtained by converting the vaccination coverage and campaign duration outcomes from the supply chain model as

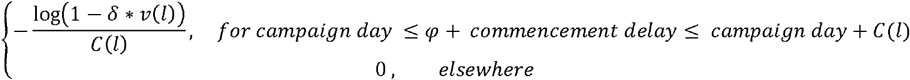

where *c*(*I*) is the campaign duration in location *I* derived in (0.7), and *φ*is the pre-deployment delay. We used *δ = 0*.*999* to slightly scale the vaccination coverage estimated from the supply chain model, to avoid the log transformation above going to infinity when *V*(*I*) *= 1*.

### Main analysis

Using the modelling framework, we simulated the 10 vaccination strategies briefly described in pairs below and summarised individually in Table S3 of the supplementary material.

1. **10-dose in full cold chain (strategy 1a & b):** Both fixed post and mobile teams use 10-dose vials transported in passive cold chain equipment (Mobile teams use RCW25s in alternative a or vaccine carriers in alternative b) with ice packs. This strategy is currently in use in most low-income settings.
2. **10-dose OCC (strategy 2a & b):** Both fixed and mobile teams use 10-dose vials transported in passive cold chain equipment (Mobile teams use RCW25s in alternative a or vaccine carriers in alternative b) without ice packs.
3. **Monodose in full cold chain (strategy 3a & b):** This strategy is like (1) except the monodose vials are used instead of 10-dose vials.
4. **Monodose OCC (strategy 4a & b):** The strategy is like (2) except the monodose vials are used instead of 10-dose vials.
5. **Partial cold chain (strategy 5a & b):** Fixed post teams use the 10-dose vials in the cold chain, but the mobile teams use the monodose vaccines OCC and transport them either in RCW25s (a) or vaccine carriers (b).

We ran simulations for each strategy, using a set of parameters from our assumptions, various literature sources, and expert opinion from our MSF collaborators (Table S4 & S5). We obtained the logistical data from the equipment manufacturers’ websites, manuals, and calculations based on the WHO’s Performance, Quality and Safety (PQS) devices catalogue [23].

We simulated a campaign where fixed post and mobile teams were dispatched sequentially to 5 locations of the same arbitrary population sizes. We used a proportional allocation scheme to allocate the two team types subject to a fixed number of teams, so that their sizes were proportional to the target population sizes.

We assumed a pre-deployment delay of 21 days to account for the time until the vaccines arrive at the field base. The choice of pre-deployment delay can influence the cases averted, so we explored its impact through a sensitivity analysis.

For simplicity, we assumed that the outbreaks in each population type and location ran the same course but independent of each other. All the simulations were initialized with the same conditions. We divided the populations at each location into near and far with sizes of 75, 000 and 25, 000, respectively. Our choice of population sizes was arbitrary.

We initialized the simulations with 75% of the total target populations, *N*, as immune, *R*(0). We assumed that this level of immunity results from previous vaccination or infection with the wildtype virus and reflects a value in the range reported by various sources [3,24–26]. We ran the model with no exposed individuals, *E*(0), and 10 initially infected individuals, *I*(0). The remainder, *N-N−E* (0) *- I*(0)*- R*(0)), were initially susceptible individuals. The population size remained constant throughout the epidemic. We ran the simulation for 365 days and aggregated the results by strategy based on the outbreak size (total cases) across all 5 locations. Moreover, we constrained the campaign to 10 days per location, similar to what is done in practice [4].

We compared the strategies across all 5 locations based on the supply chain results of campaign duration and vaccination coverage and the cases averted from the epidemiological model results. To obtain the cases averted, we calculated the total outbreak sizes from each strategy across all 5 locations and defined the cases averted as the difference between the total outbreak size of each strategy and strategy 1b (Table S3), which served as the baseline.

### Sensitivity analysis

We conducted two sensitivity analyses. First, we analysed the impact of different sizes of near and far populations, and team allocation schemes at each location on vaccination coverage and campaign duration. We kept every other assumption the same as in the main analysis. The two team allocation schemes we considered were: *proportional*, where the total teams were allocated to the two team types proportional to the target population type sizes, and *equal*, where the total teams were allocated to the two team types in an equal split irrespective of the target population size. We allocated the near and far populations at each location so that for a campaign targeting 5 locations each with total population of 100 000, the target population sizes in terms of near and far were in ratios of 1:4, 2:3, 2.5:2.5, 3:2, and 4:1, respectively. To present the results, we provide the mean vaccination coverage for the two team allocation schemes and across all strategies by each near and far population size scenario.

Second, we explored the impact of various pre-deployment delays on the cases averted. We varied pre-deployment delays from 21-84 days in steps of 7 days and fixed every other parameter value as in the main analysis.

## Results

### Main analysis

In the main analysis (Table S4), all strategies, except the monodose in full cold chain with vaccine carrier as the mobile team equipment, achieved high coverage of approximately 80% (Fig. 3). The OCC, partial cold chain, and full cold chain strategies took 50, 51, and 52 days to complete respectively, regardless of the transport equipment used (Fig. 3).

**Fig. 3.**
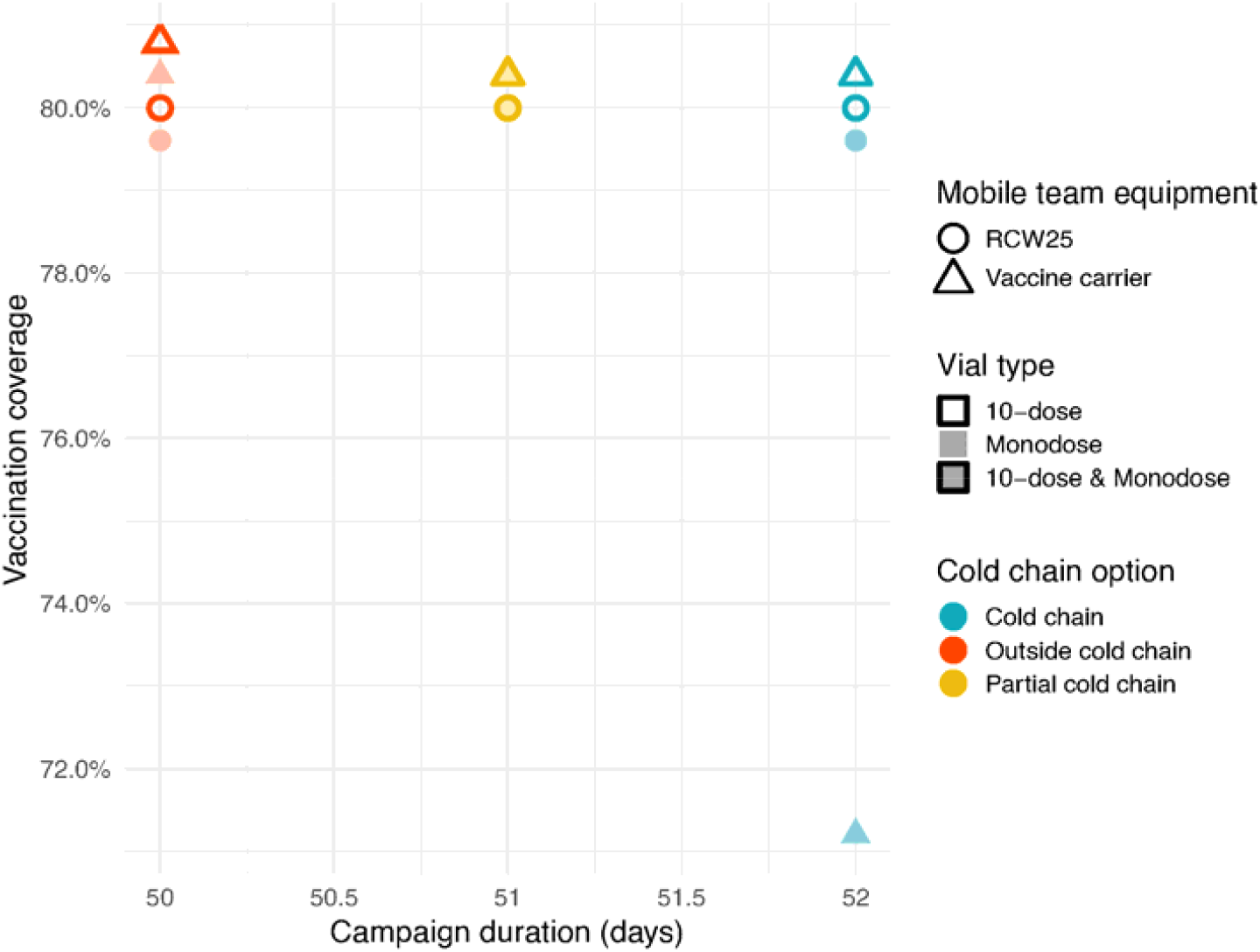
Ranking of the strategies based on the vaccination coverage and campaign duration at the end of the campaign. Cold chain strategies are illustrated in turquoise, partial cold chain strategies in gold, and OCC strategies in orange. Filled shapes represent strategies with the monodose vials only. Shapes with a stroke but no fill represent strategies using the 10-dose. Mixed strategies are represented by shapes with both fill and stroke. The circles and triangles represent respectively the strategies with mobile teams using RCW25 and vaccine carrier as transport equipment. Points of the same (x, y) position are shifted vertically to prevent overlap but have the same value.

All the full cold chain strategies resulted in either the same a or fewer cases averted than the baseline strategy of 10-dose vials in full cold chain, using vaccine carriers for the mobile teams (Strategy 1b in Table S3). The partial cold chain strategies averted more cases than the baseline, but the OCC strategies averted the most cases relative to the baseline (Fig. 4).

**Fig. 4.**
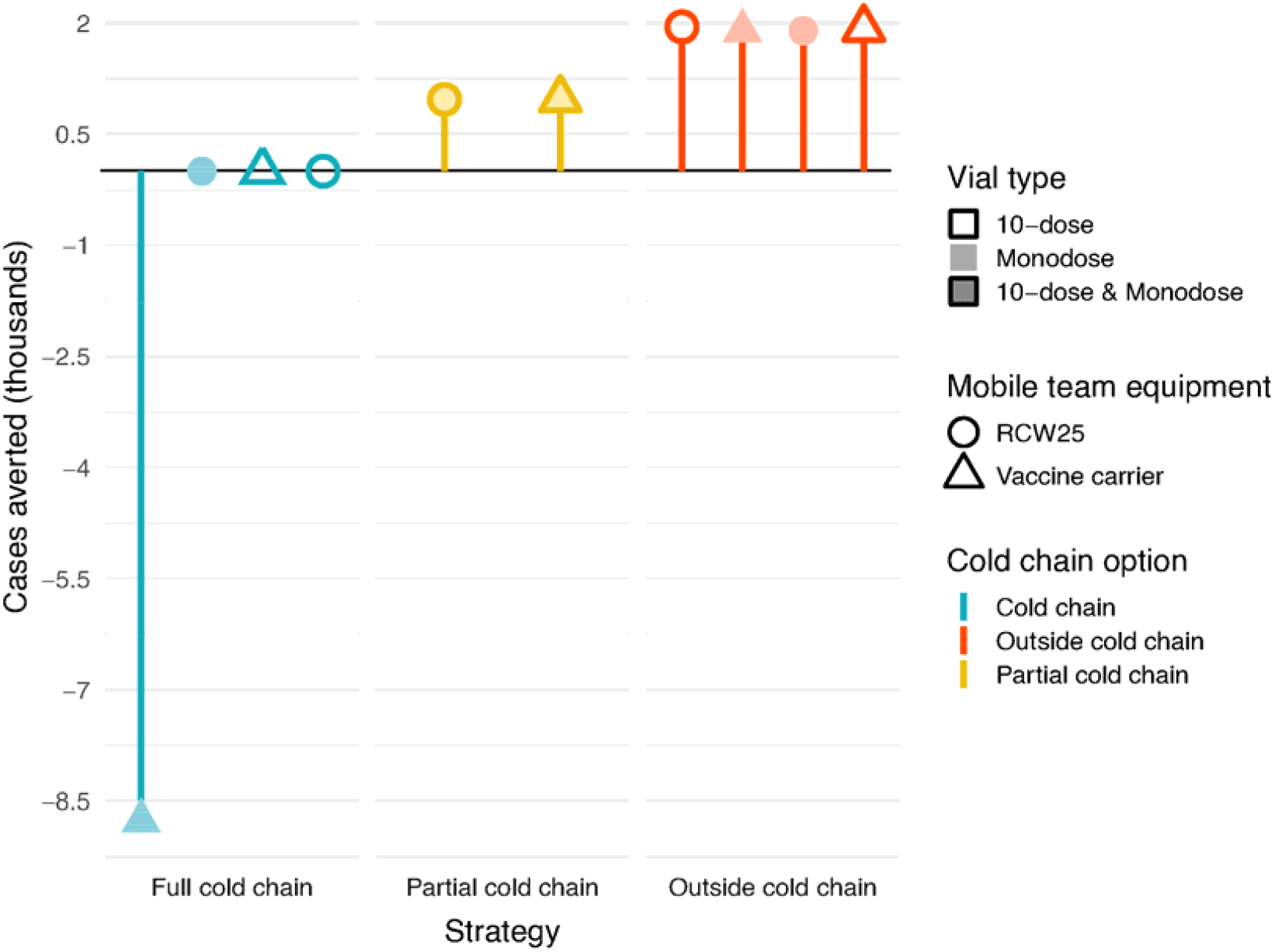
Cases averted by strategy. To calculate the cases averted, we chose as the baseline the 10-dose in the full cold chain strategy that assumes that the mobile teams use vaccine carrier as the transport equipment (turquoise open triangle). Cold chain strategies are represented in turquoise, partial cold chain strategies in gold, and OCC strategies in orange. Filled shapes represent strategies with the monodose vials only. Shapes with a stroke but no fill represent strategies using 10-dose vials. Mixed strategies are represented by shapes with both fill and stroke. The circles and triangles represent respectively the strategies with mobile teams using RCW25 and vaccine carrier as transport equipment.

### Sensitivity analyses

#### Impact of varying population sizes and team allocation schemes on vaccination coverage and duration

The proportional team allocation scheme always leads to a higher or equal vaccination coverage relative to the equal team allocation and scales monotonically as the proportion of near to far population increases (Fig. 5). The equal team allocation results in proportionately lower coverage when the near population is either small or large.

**Fig. 5.**
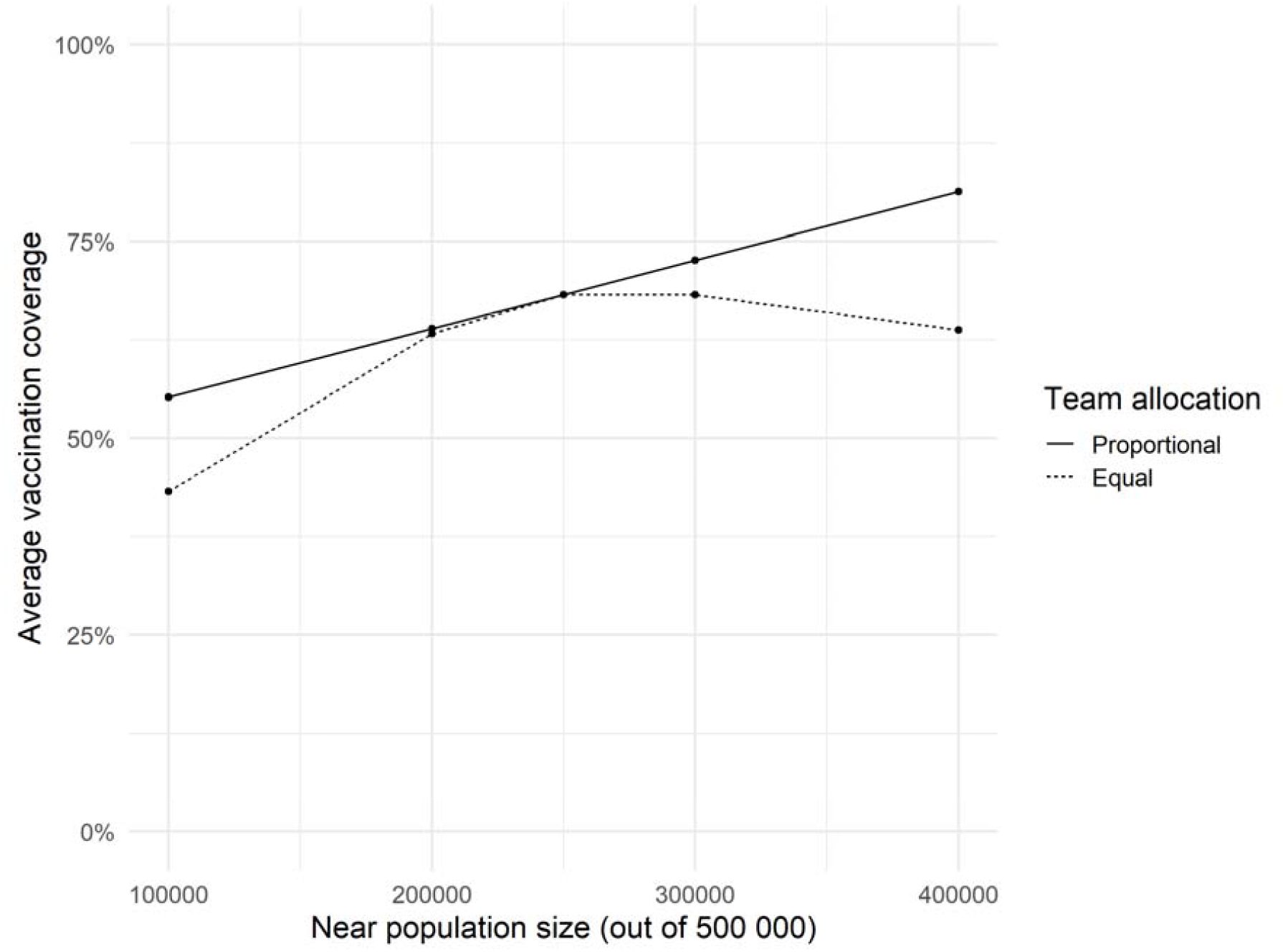
Average vaccination coverage across strategies for proportional and equal team allocation schemes with varying population sizes. Each point was obtained by averaging over the vaccination coverage obtained across all the strategies for each near to far population ratio and team allocation scheme. The solid line represents the proportional team allocation scheme, and the dashed line represents the equal team allocation.

#### Impact of pre-deployment delay on the cases averted

The OCC strategies avert the most cases when there is a short pre-deployment delay (Fig. 6). The partial cold chain and full cold chain strategies follow respectively in terms of the number of the cases averted, except the monodose full cold chain with vaccine carrier as mobile team transport equipment. The difference in cases averted between the strategies, however, decreases as the delay increases. The strategies at this time point are similar in terms of cases averted. These results show a clear relationship between the delay before a campaign is deployed and the cases that could potentially be averted.

**Fig. 6.**
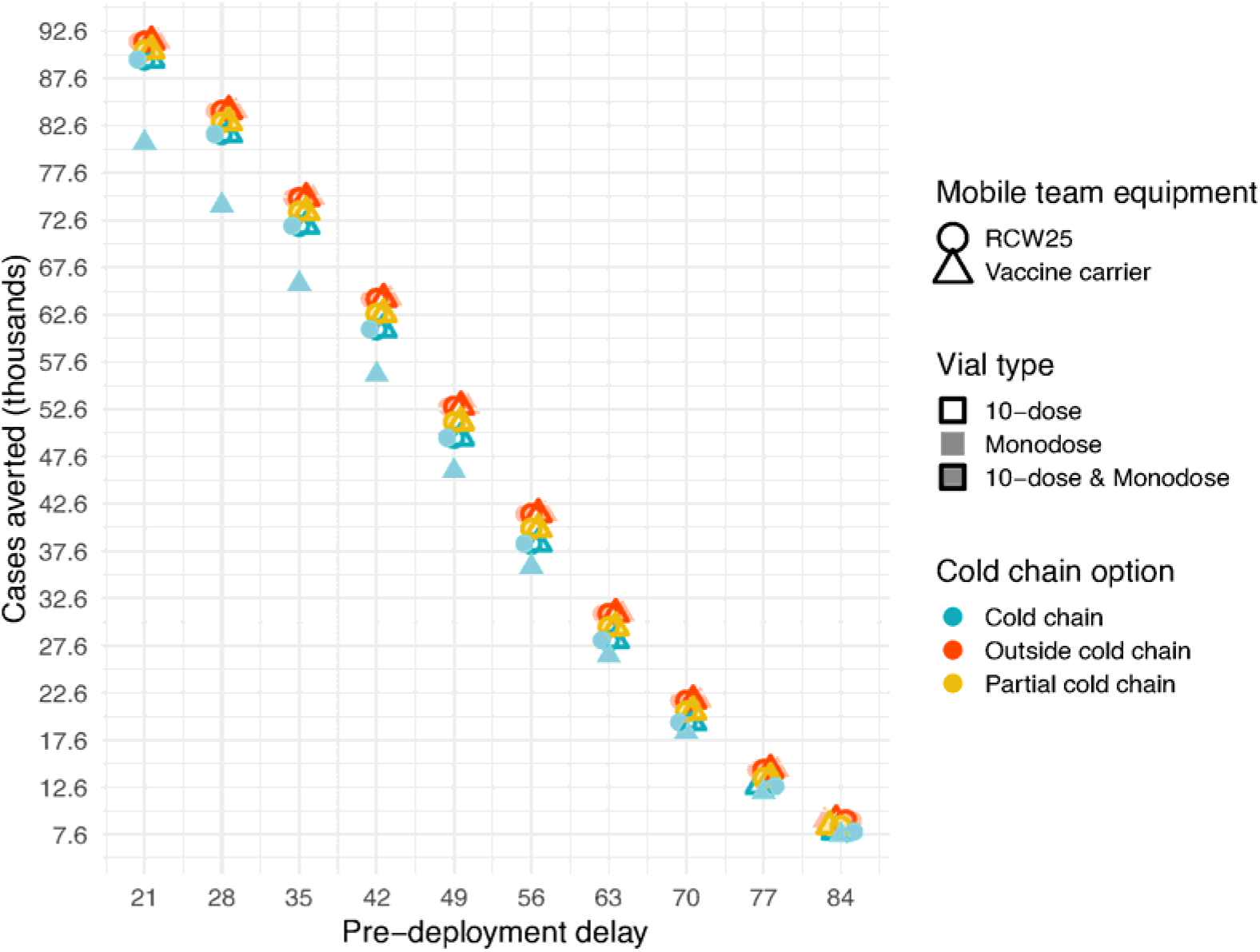
Impact of pre-deployment delay on cases averted. Strategies are ranked based on the cases averted. Cold chain strategies are illustrated in turquoise, partial cold chain strategies in gold, and OCC strategies in orange. Filled shapes represent strategies with the monodose vials only. Shapes with a stroke but no fill represent strategies using the 10-dose. Mixed strategies are represented by shapes with both fill and stroke. The circles and triangles represent respectively the strategies with mobile teams using RCW25 and vaccine carrier as transport equipment. Points of the same (x, y) position are shifted vertically to prevent overlap but have the same value.

## Discussion

We have found that the OCC strategies could result in campaigns with shorter durations and a greater number of cases averted. The shorter campaign duration is because there is no initial delay in campaign commencement due to the elimination of the need for ice packs to transport the vaccines in the last mile of the campaign. During measles outbreak response vaccination, a quick response, assuming no loss of vaccination coverage, is always preferable in minimizing outbreak size [24,27–31].

We have developed a modelling framework for evaluating the potential use of the measles vaccine OCC for outbreak response. The framework allows us to compare alternative strategies based on vaccination coverage, campaign duration, and cases averted. We estimated the time it takes to freeze ice packs prior to dispatching vaccination teams for a campaign. We used this to the approximate the delay to commence a campaign requiring the cold chain. Moreover, the framework calculates the total operational time to complete a campaign (campaign duration) targeting several locations with a given number of teams, and the expected vaccination coverage. These outcomes are natural inputs to an epidemiological model for simulating transmission dynamics to quantify the number of cases averted through a series of intermediate steps.

We have observed a clear rank of the strategies in our analysis of the supply chain and epidemiological outcomes. We have found that the OCC strategies are always best in terms of campaign duration and cases averted, followed by the partial cold chain strategies, and the full cold chain strategies. All but one of the strategies achieve the same high vaccination coverage (80%), but the absolute value of coverage is subject to our assumptions. Therefore, if the choice is to use the vaccines outside of cold chain, it does not matter what passive cold chain equipment the mobile teams use since, not needing ice packs, they all allow for enough dose transport capacity at no cost of time.

The benefits of the OCC strategies are subject to several logistical challenges including pre-deployment delays, team allocation schemes, and team performance. Even though the strategy could potentially avert more cases due to reduced delays, the magnitude of the pre-deployment delay could diminish the benefit of averting more cases (Fig. 6). Moreover, the strategy’s potential vaccination coverage depends on the team allocation scheme used. When the total teams available are allocated to the fixed post and mobile teams in proportion to the size of the near and far populations, the strategy yields a much higher coverage than when the teams are allocated equally regardless of the target population. The potential benefits of the OCC strategy therefore can only be fully harnessed by implementing it rapidly and with the appropriate team allocation scheme.

We used four phenomena to synthesize the factors that affect the success of a campaign: pre-deployment delays, campaign commencement delays, the time it takes to complete a campaign (campaign duration), and vaccination coverage. When a measles outbreak is ongoing, the susceptible population diminishes quite quickly in the absence of an intervention. Hence, for a strategy with the potential to achieve a high coverage, if the campaign starts late when there are not as many susceptible individuals to be vaccinated, the full potential of the campaign might not be achieved. Secondly, given that most outbreak response activities are constrained to a limited time in each location, the time it takes to complete a campaign must be used efficiently by channelling adequate resources into the campaign. This can be achieved, for example, by dispatching the needed teams to complete the campaign within the given campaign duration. This was apparent in our sensitivity analysis that examined dispatching teams in proportion to the population size (Fig. 5). The potentially high coverage expected of a strategy can be hindered if the logistical needs, for example teams and transport equipment, for achieving such a high coverage are not met. This may explain the low coverages stated in the literature [4].

We have identified the operational constraints associated with the monodose and 10-dose vials in and outside cold chain. We have found that the large volume per dose of the monodose vial means it is never optimal for use with the conventional vaccine carrier, especially in the cold chain. If a larger carrier (example, an RCW25 cold box) is used, then both the monodose and 10-dose strategies can carry enough doses to meet the expected team performance and are thus equivalent.

The decision to use OCC strategies should be accompanied by a thorough understanding of how many vaccines need to be transported on each day of the campaign. Dispatched teams will have to transport enough doses to achieve the team performance while simultaneously minimizing how many vaccines are exposed to ambient temperature. Vaccines that are exposed for longer than the specified duration will have to be discarded, counting as wastage, and could impact the cost of campaigns. Future work will employ other methods to capture the uncertainties in team performance to understand the interplay between vaccine usage and wastage during campaigns and how it affects the outcomes studied here.

This study has several limitations. We assumed that the populations mix homogenously. Population mixing patterns are often heterogeneous and complex and our assumption was made for the sake of simplicity. We also assumed that the locations had the same total population sizes and experienced independent epidemics but with the same underlying dynamics, starting with the same initial population structure. Epidemics in different locations are never the same in every respect. Additionally, the model did not account for population migration and movement between the locations. We know that populations are often interconnected and therefore have some possibility of importing infections [32].

We also defined the open vial wastage and team performance as inputs to the model, meaning that the teams know ahead of time the population sizes that will turn out for vaccination and hence how many vaccines to transport. This assumption could be modified, for example by treating the population sizes presenting for vaccination as a stochastic variable.

We also assumed that the campaign would occur sequentially across locations, but in some campaigns, teams are dispatched in parallel to several locations at the same time.

All these limitations reflect the fact that models are an abstraction of real phenomena and do not reflect the exact occurrences. Therefore, our findings aim to aid in decision-making but do not represent exact expectations of any future campaigns being planned with measles vaccines using an OCC strategy.

### Conclusion

Conducting measles outbreak response campaigns in the cold chain in low-income countries requires dealing with many logistical challenges. OCC campaigns have the potential to speed up measles outbreak response and reduce the time it takes to complete campaigns, achieving high vaccination coverage, and averting a high number of cases. The benefits presented by this alternative strategy could enhance the control and elimination of measles. We therefore recommend that the various vaccine stakeholders consider prioritising the licensure, prequalification, and relabelling of the appropriate measles vaccines for OCC use under ECTC.

## Data Availability

All data generated in the analysis are shared in the manuscript.

## CRediT authorship contribution statement

**James M Azam:** Conceptualization, Methodology, Software, Visualization, Formal analysis, Writing – original draft, Writing – review and editing. **Kimberly Bonner:** Conceptualization, Writing – review and editing. **Barbara Saitta:** Conceptualization, Writing – review and editing. **Matthew J Ferrari:** Conceptualization, Methodology, Supervision, Writing – review and editing. **Juliet RC Pulliam:** Conceptualization, Methodology, Supervision, Writing – review and editing.

## Acknowledgements

We would like to thank the members of the MSF Vaccine Working Group, who continuously availed themselves to help conceptualize and improve this work with their invaluable experience and feedback.

## Funding

This work is based on the research supported by the Department of Science and Innovation and the National Research Foundation. Any opinion, finding, and conclusion or recommendation expressed in this material is that of the authors and the NRF does not accept any liability in this regard.

## Declaration of Competing Interest

The authors declare no competing interests.

## Supplementary materials

### Methods

Below are the tables referred to in the main article.

**Table S1.** MF314 freezer specification, ice pack holding capacity, and ice pack freezing rates.

| Item | Freezing rate (ice packs per day) | Capacity |
| --- | --- | --- |
| Large ice packs (0.6 L) | 54 | 323 |
| Small ice packs (0.4 L) | 81 | 450 |

**Table S2.**
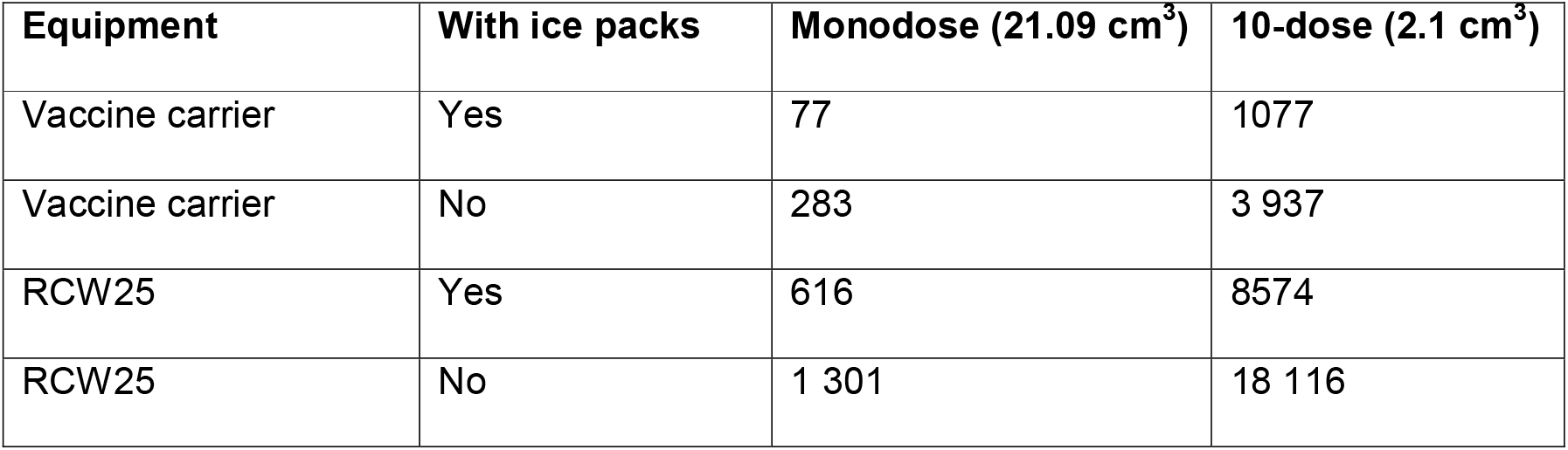
Passive cold chain equipment capacities for monodose and 10-dose with and without ice packs. For both vial types, we calculated the capacities without ice by dividing the total internal volume by the packed volume per dose. For the capacities with ice packs, we divided the equipment’s net volume after ice packs by the packed volume per dose. We obtained the equipment information from the manufacturer’s website and the WHO’s Performance, Quality and Safety (PQS) devices catalogue.

| Equipment | With ice packs | Monodose (21.09 cm<sup>3</sup>) | 10-dose (2.1 cm<sup>3</sup>) |
| --- | --- | --- | --- |
| Vaccine carrier | Yes | 77 | 1077 |
| Vaccine carrier | No | 283 | 3 937 |
| RCW25 | Yes | 616 | 8574 |
| RCW25 | No | 1 301 | 18 116 |

**Table S3.**
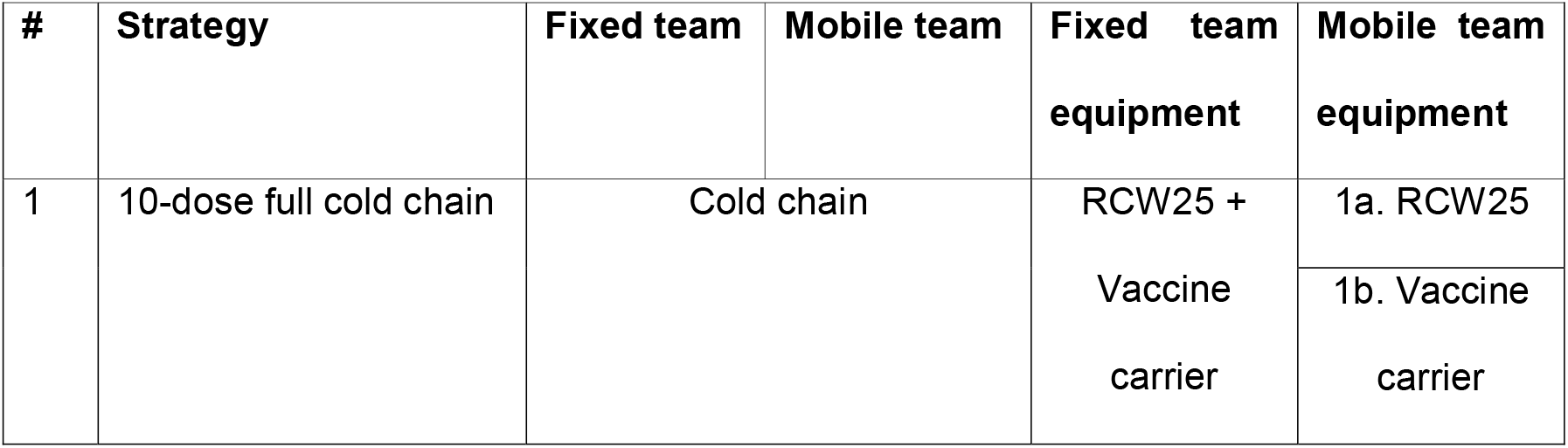
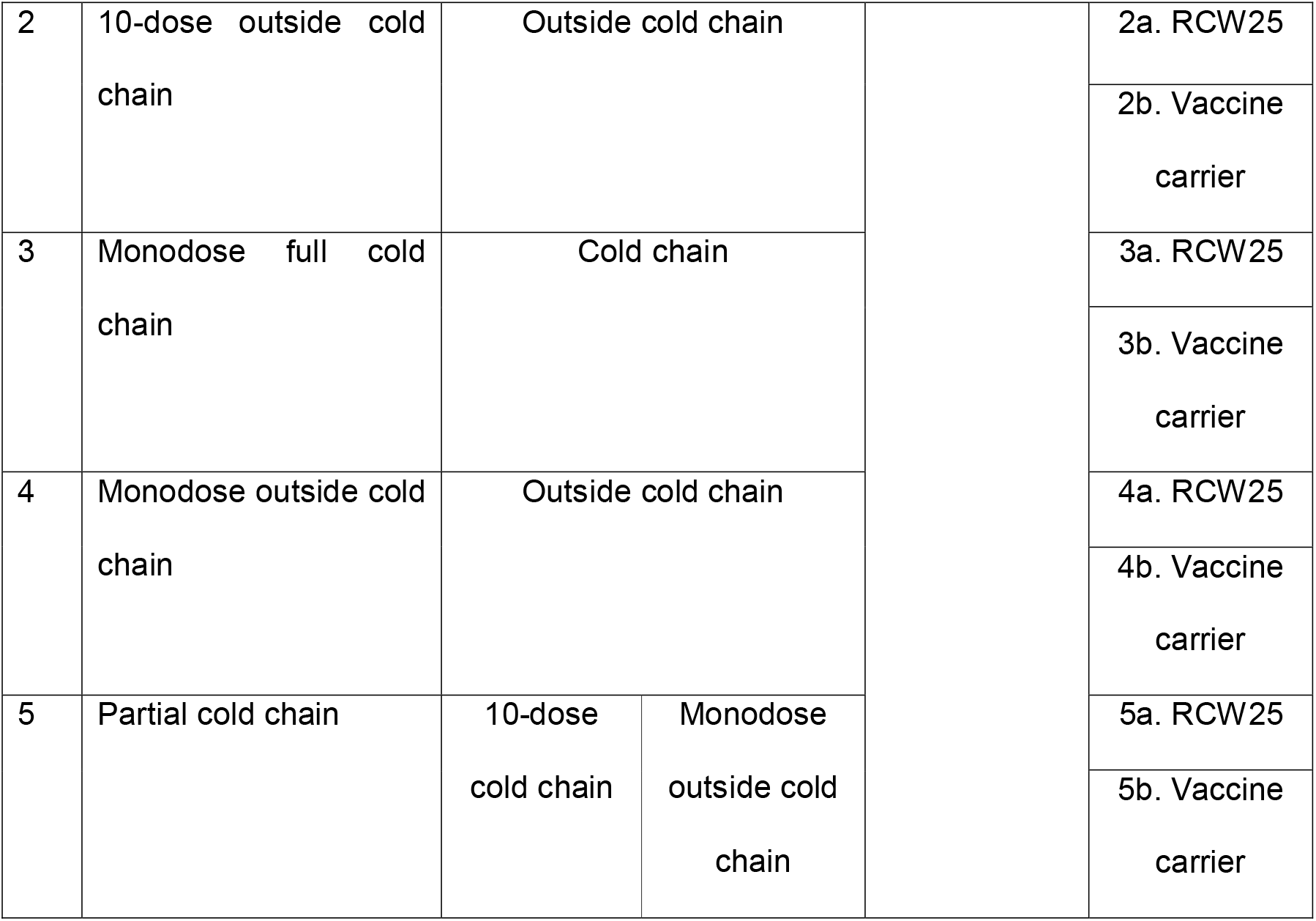
Summary of the composition of each strategy in terms of the vial type and cold chain options for the fixed and mobile teams. For each strategy, we assumed the fixed post teams used both the RCW25 and vaccine carrier at the fixed post. We assumed there were two fixed post teams per site so that they used 1 RCW25 and 2 vaccine carriers. For each strategy, we considered the option of mobile teams either using only an RCW25 or a vaccine carrier so that in all, we simulated 10 strategies for comparison.

**Table S4.** Supply chain simulation parameters for the main analysis.

| Description | Value/Range | Source |
| --- | --- | --- |
| Number of MF314 freezers | 10 | Assumed |
| Number of locations | 5 | Assumed |
| Near population per location | 75 000 | Assumed |
| Far population per location | 25 000 | Assumed |
| Number of fixed post teams | 10 | Assumed |
| Number of mobile teams | 10 | Assumed |
| Fixed posts per vaccination site | 2 | [1] |
| Maximum campaign duration per location | 10 days | MSF |
| Pre-deployment delay | 21-84 days (in 7-day steps) | Assumed |
| Team performance (fixed team) | 450 people per day | [1] |
| Team performance (mobile team) | 250 people per day | [1] |
| Buffer stock | 15% | [1] |
| Open vial wastage 10-dose (mobile team) | 15% | [1] |
| Open vial wastage 10-dose (fixed post team) | 15% | [1] |
| Open vial wastage monodose (mobile team) | 3% | MSF |
| Open vial wastage monodose (fixed post team) | 3% | MSF |
| Number of days to replace large ice packs | 2 | [1] |
| Number of days to replace small ice packs | 1 | [1] |

**Table S5.** Epidemiological model simulation parameters.

| Parameter | Description | Value | Source |
| --- | --- | --- | --- |
| $R_0$ | Basic reproduction number | 12.8 | [2] |
| $v$ | Vaccine efficacy | 0.84 | [3] |
| $1/r$ | Latency period | 10 days | [4] |
| $s1/g$ | Infectious period | 8 days | [4] |

## Notes

### Competing Interest Statement

The authors have declared no competing interest.

### Author Declarations

This research does not need ethical approval.

## References

[1] Roberts L. Why measles deaths are surging — and coronavirus could make it worse. Nature 2020;580:446–7. doi:10.1038/d41586-020-01011-6.

[2] World Health Organization, WHO, World Health Organization. Measles vaccines: WHO position paper, April 2017 - Recommendations. Vaccine 2017;37:205–27. doi:10.1016/j.vaccine.2017.07.066.

[3] Keating P, Isidro A, Martin C, Blake A, Lechevalier P, Uzzeni F, et al. Measles seroprevalence after reactive vaccination campaigns during the 2015 measles outbreak in four health zones of the former Katanga Province, Democratic Republic of Congo 2019:1–11.

[4] Gerard SP, Kyrousis E, Zachariah R. Measles in the Democratic Republic of the Congo: an urgent wake-up call to adapt vaccination implementation strategies. Public Heal Action 2014;4:6–8.

[5] Juan-Giner A, Alsalhani A, Panunzi I, Lambert V, Van Herp M, Gairola S. Evaluation of the stability of measles vaccine out of the cold chain under extended controlled temperature conditions. Vaccine 2020;38:2473–7. doi:10.1016/j.vaccine.2020.02.005.

[6] WHO. Guidelines on the stability evaluation of vaccines for use under extended controlled temperature conditions. Guidance 2015:2268. doi:WHO/BS/2015.2268.

[7] WHO. How Do Ctc and Ectc Relate To Each Other n.d.:40.

[8] Juan A, Panunzi I, Alsalhani A, Lambert V, Barrie I, Herp M Van. Evaluation of the stability of the measles vaccine for use in Extended Controlled Temperature Conditions (ECTC) 2018.

[9] Zipursky S, Djingarey MH, Lodjo JC, Olodo L, Tiendrebeogo S, Ronveaux O. Benefits of using vaccines out of the cold chain: Delivering Meningitis A vaccine in a controlled temperature chain during the mass immunization campaign in Benin. Vaccine 2014;32:1431–5. doi:10.1016/j.vaccine.2014.01.038.

[10] Lydon P, Zipursky S, Tevi-Benissan C, Djingarey MH, Gbedonou P, Youssouf BO, et al. Economic benefits of keeping vaccines at ambient temperature during mass vaccination: the case of meningitis A vaccine in Chad. Bull World Health Organ 2014;92:86–92. doi:10.2471/BLT.13.123471.

[11] Ramirez-Gonzalez A, Hennessey K, Vilayvone V, Xeuatvongsa A, Vongxay V, Patel MK, et al. Hepatitis B vaccine stored outside the cold chain setting: a pilot study in rural Lao PDR. Vaccine 2016;34:3324–30. doi:10.1016/j.vaccine.2016.03.080.

[12] Patel MK, Kahn A. Comment Game changing L: hepatitis B vaccine in a controlled temperature chain. Lancet Glob Heal n.d.;6:e596.-e597. doi:10.1016/S2214-109X(18)30233-X.

[13] World Health Organization, Controlled Temperature Chain Working Group. Controlled Temperature Chain: Strategic Roadmap for Priority Vaccines 2017-2020. Who/Ivb/1720 2017.

[14] Dadari IK, Zgibor JC. How the use of vaccines outside the cold chain or in controlled temperature chain contributes to improving immunization coverage in low-and middle-income countries (LMICs): A scoping review of the literature. J Glob Health 2021;11:1–14. doi:10.7189/jogh.11.04004.

[15] Grais RF, Conlan AJK, Ferrari MJ, Djibo A, Le Menach A, Bjornstad ON, et al. Time is of the essence: exploring a measles outbreak response vaccination in Niamey, Niger. J R Soc INTERFACE 2008;5:67–74. doi:10.1098/rsif.2007.1038.

[16] Li S, Ferrari MJ, Bjørnstad ON, Runge MC, Fonnesbeck CJ, Tildesley MJ, et al. Concurrent assessment of epidemiological and operational uncertainties for optimal outbreak control: Ebola as a case study. Proc R Soc B Biol Sci 2019;286:20190774. doi:10.1098/rspb.2019.0774.

[17] Shea K, Tildesley MJ, Runge MC, Fonnesbeck CJ, Ferrari MJ. Adaptive Management and the Value of Information: Learning Via Intervention in Epidemiology. PLoS Biol 2014;12:9–12. doi:10.1371/journal.pbio.1001970.

[18] Comes T, Bergtora Sandvik K, Van de Walle B. Cold chains, interrupted: The use of technology and information for decisions that keep humanitarian vaccines cool. J Humanit Logist Supply Chain Manag 2018;8:49–69. doi:10.1108/JHLSCM-03-2017-0006.

[19] Lee BY, Norman BA, Assi TM, Chen SI, Bailey RR, Rajgopal J, et al. Single versus multi-dose vaccine vials: An economic computational model. Vaccine 2010;28:5292– 300. doi:10.1016/j.vaccine.2010.05.048.

[20] Guichard S, Hymbaugh K, Burkholder B, Diorditsa S, Navarro C, Ahmed S, et al. Vaccine wastage in Bangladesh. Vaccine 2010;28:858–63. doi:10.1016/j.vaccine.2009.08.035.

[21] Danet C, Fermon F. Management of a measles epidemic - a practical guide for doctors, nurses, laboratory technicians, medical auxilliaries and logisticians. Medicins Sans Frontieres; 2013.

[22] Bjørnstad ON. Epidemics. Model Data Using R Springer Int Publ 2018:318. doi:https://doi.org/10.1007/978-3-319-97487-3.

[23] WHO. PQS devices catalogue. Who/Ivb/1108 2017:1–392.

[24] Grais RF, Conlan AJK, Ferrari MJ, Djibo A, Le Menach A, Bjørnstad ON, et al. Time is of the essence: Exploring a measles outbreak response vaccination in Niamey, Niger. J R Soc Interface 2008;5:67–74. doi:10.1098/rsif.2007.1038.

[25] Winter AK, Wesolowski AP, Mensah KJ, Bruna M, Randriamanantena AH, Cauchemez S, et al. Revealing Measles Outbreak Risk with a Nested IgG Serosurvey in Madagascar 2018. doi:10.1093/aje/kwy114/5033615.

[26] Trentini F, Poletti P, Merler S, Melegaro A. Measles immunity gaps and the progress towards elimination: a multi-country modelling analysis. Lancet Infect Dis 2017;17:1089–97. doi:10.1016/S1473-3099(17)30421-8.

[27] Duijzer LE, van Jaarsveld W, Dekker R. The benefits of combining early aspecific vaccination with later specific vaccination. Eur J Oper Res 2018;271:606–19. doi:10.1016/j.ejor.2018.05.054.

[28] Ferrari MJ, Fermon F, Nackers F, Llosa A, Magone C, Grais RF. Time is (still) of the essence: quantifying the impact of emergency meningitis vaccination response in Katsina State, Nigeria. Int Health 2014;6:282–90. doi:10.1093/inthealth/ihu062.

[29] Grais RF, Conlan AJK, Ferrari MJ, Djibo A, Le Menach A, Bjørnstad ON, et al. Time is of the essence: exploring a measles outbreak response vaccination in Niamey, Niger. J R Soc Interface 2008;5:67–74. doi:10.1098/rsif.2007.1038.

[30] Ferrari MJ, Fermon F, Nackers F, Llosa A, Magone C, Grais RF. Time is (still) of the essence: Quantifying the impact of emergency meningitis vaccination response in Katsina State, Nigeria. Int Health 2014;6:282–90. doi:10.1093/inthealth/ihu062.

[31] Grais RF, D. Radiguès X, Dubray C, Fermon F, Guerin PJ, Fermonn F, et al. Exploring the time to intervene with a reactive mass vaccination campaign in measles epidemics. Epidemiol Infect 2006;134:845–9. doi:10.1017/S0950268805005716.

[32] Ferrari MJ, Grais RF, Bharti N, Conlan AJK, Bjørnstad ON, Wolfson LJ, et al. The dynamics of measles in sub-Saharan Africa. Nature 2008;451:679–84.

## References

[1] Danet C, Fermon F. Management of a measles epidemic - a practical guide for doctors, nurses, laboratory technicians, medical auxilliaries and logisticians. Medicins Sans Frontieres; 2013.

[2] Guerra FM, Bolotin S, Lim G, Heffernan J, Deeks SL, Li Y, et al. The basic reproduction number (R0) of measles: a systematic review. Lancet Infect Dis 2017. doi:10.1016/S1473-3099(17)30307-9.

[3] World Health Organization, WHO, World Health Organization. Measles vaccines: WHO position paper, April 2017 - Recommendations. Vaccine 2017;37:205–27. doi:10.1016/j.vaccine.2017.07.066.

[4] Heymann DL. Control of communicable diseases manual 18th Edition 2004:417.

